# Anxiolytic management in eating disorder patients receiving cognitive behavioral therapy: A quality improvement brief report

**DOI:** 10.1101/2024.11.25.24317936

**Authors:** Brad E.R. Smith, Katelyn A. Greenberg, Sreya Vadapalli, Sheldon R. Garrison

## Abstract

Eating disorders (EDs) are serious lifelong and debilitating health conditions characterized by complex adverse eating behaviors resulting in weight loss and decreased quality of life. One evidence-based treatment is a form of cognitive-behavioral therapy (CBT) called exposure therapy where patients progress through a hierachy of exposures. This treatment approach results in concern related to CBT efficacy for patients concurrently treated with anxiolytic medications. These medications, which are being incorporated into treatment protocols, may impact the anxiety-provoking components of the exposure response, potentially limiting its efficacy. Therefore, patients were assessed in multiple ways to ensure that providers could prescribe anxiolytics without risk to diminishing the well-established response to CBT. To ensure that patients treated with anxiolytics progressed at a similar rate to those who did not receive them, a quality improvement study was conducted to compare all patients in residential, partial hospitalization and intensive outpatient eating disorder treatment programs.

## Introduction

The impact of eating disorders (EDs) is felt broadly, with a lifetime prevalence in the general population of 0.9% (Qian et al., 2022). The onset of EDs, most commonly anorexia nervosa, bulimia nervosa and binge eating disorder, begins with behavioral and cognitive abnormalities, leading to multi-system issues. Early symptoms may manifest as difficult eating under 10 years (Pinhas et al., 2011), transitioning in adolescence to dieting, body dissatisfaction and body image concerns during critical neurodevelopmental periods (Rohde et al., 2015).

The complex neurocognitive involvement of EDs results in inadequate pharmacological options. Current evidence-based treatment for EDs is cognitive behavioral therapy (CBT), specifically with fear extinction and anti-depressants (Crone et al., 2023; Farrell et al., 2019; P. Hay, 2020). Comorbid disorders present in 56-92% of ED patients, including over half with anxiety (Wade et al., 2023). This adds an unexpected complexity to treating when anxiolytic medications are implemented to manage anxiety, introducing concerns regarding how they may affect CBT efficacy.

The goal of this retrospective quality improvement study was to analyze changes in ED symptom severity, as well as severity of co-occurring depressive and obsessive-compulsive disorder (OCD) symptoms in a high-acuity patient population undergoing ED recovery. Providers began incorporating anxiolytics for the management of patients with an ED. To ensure the anxiolytics did not interfere with other treatment modalities, patients prescribed anxiolytics were evaluated using multiple assessments during the course of their treatment. Improvement for patients receiving anxiolytic treatment (anxiolytic) with standard of care was compared to those receiving standard of care alone (non-anxiolytic) to ensure a consistent degree of improvement.

## Methods

Electronic medical records were retrospectively analyzed for all patients discharged from an ED program at Rogers Behavioral Health, a behavioral healthcare system that provides mental health and addiction treatment services, between January 1, 2018 and May 31, 2024. Inclusion criteria were: (1) ≥18 years of age; (2) completion of pre- and post-assessments given at admission (baseline); and, at discharge for at least one measure; (3) admission to an ED program at any treatment level (inpatient, residential, partial hospitalization and intensive outpatient). Each patient’s most intensive level of care was analyzed. All patients received intensive daily cognitive-behavioral therapy (CBT) with exposure and response prevention (ERP), a standard of care treatment option. The anxiolytics administered to patients were prescribed according to physician preference and clinical judgment, and included alprazolam, clonazepam, diazepam, gabapentin, hydroxyzine, and lorazepam. Patient data was extracted from the electronic health records and de-identified.

### Outcomes

The primary outcome measure was the Eating Disorder Examination Questionnaire (EDE-Q) (Fairburn & Beglin, 1994) to measure ED behavior frequency from admission to discharge. Secondary outcomes included the Yale-Brown Cornell Eating Disorder Scale (YBC-EDS) (Mazure et al., 1994) to measure ED symptom severity, the Quick Inventory of Depression Symptomatology (QIDS) scale (Rush et al., 2003) to measure depressive symptoms and the Yale-Brown Obsessive-Compulsive Scale (Y-BOCS) scale to measure obsessive-compulsive disorder (OCD) symptom severity (Goodman, Price, Rasmussen, Mazure, Delgado, et al., 1989; Goodman, Price, Rasmussen, Mazure, Fleischmann, et al., 1989). Additional secondary outcomes included exposure hierarchy percent completion to quantify CBT treatment progress, length of stay (LOS), and readmission rate. The demographic variables collected included age, sex, race, and ethnicity. An additional clinical variable employed in the analysis was level of care, consisting of inpatient, residential, partial hospitalization, and intensive outpatient.

### Data analysis

The treatment groups for the following analyses were divided into those who were prescribed anxiolytic medication alongside standard therapy (‘anxiolytic’) and those not prescribed anxiolytics (‘non-anxiolytic’). The non-anxiolytic group was control-matched to the anxiolytic group by randomly selecting the matched controls using the MatchIt package within R, version 4.3.3 (Ho et al., 2011; R Core Team, 2021). They were control-matched based on age, sex, race, and EDE-Q score at admission to ensure similar baseline characteristics between the anxiolytic and non-anxiolytic groups. Demographic characteristics were reported as frequencies and compared between treatment groups using Chi-square tests. Primary analyses were within- and between-group comparisons in assessment scores at admission and discharge, obtained using t tests. Assessment outcomes were reported as mean scores and standard error. Secondary analyses compared outcomes between levels of care in addition to treatment group, completed using t tests. Readmission rates were reported as frequencies and LOS was reported as number of days. For inpatient and residential, LOS is measured in calendar days, whereas partial hospitalization and intensive outpatient are measured in treatment days. Statistical analyses were completed using R.

## RESULTS

### Demographics and baseline clinical characteristics

Demographic data for the 242 patients who met eligibility criteria were reported at baseline (**Table 1**). Upon admission, EDEQ scores for both treatment groups fell within the clinical range denoting ‘moderate’ severity. YBC-EDS scores for patients taking anxiolytic medication alongside standard therapy were slightly higher than those not taking anxiolytics; the anxiolytic group fell within the clinical range indicating ‘moderate’ severity, while the non-anxiolytic group fell within the range indicating ‘mild’ severity. YBOCS-SR and QIDS scores were moderate in both groups (**Table 1**).

**Table 1.** Demographic characteristics.

|  | Anxiolytic | Non-anxiolytic | *p* value |
| --- | --- | --- | --- |
| n | 121 | 121 |  |
| Age (y) [mean (SEM)] | 25.4 (0.9) | 24.7 (0.8) | 0.6 |
| Sex (%) |  |  | 0.509 |
| Female | 83.5 | 79.3 |  |
| Male | 16.5 | 20.7 |  |
| Race and Ethnicity (%) |  |  | 0.545 |
| White | 93.4 | 88.4 |  |
| Black | 0.8 | 0.8 |  |
| Asian or Pacific Islander | 2.5 | 1.7 |  |
| Other | 1.7 | 3.3 |  |
| Ethnicity (%) |  |  | 0.418 |
| Hispanic or Latino | 7.4 | 7.4 |  |
| Not Hispanic or Latino | 91.7 | 85.1 |  |

**Treatment response, comparison between groups at baseline and discharge.**
| Variable [mean (SEM), n] | Baseline |  |  | Discharge |  |  |
| --- | --- | --- | --- | --- | --- | --- |
|  | Anxiolytic | Non-anxiolytic | *p* value | Anxiolytic | Non-anxiolytic | *p* value |
| EDEQ total score | 3.3 (0.15), 121 | 3.2 (0.1), 121 | 0.689 | 2.0 (0.1), 120 | 1.8 (0.1), 121 | 0.179 |
| YBC-EDS total score | 16.5 (0.6), 120 | 15.0 (0.6), 115 | 0.698 | 10.5 (0.6), 120 | 8.9 (0.5), 121 | 0.098 |
| QIDS total score | 14.8 (0.5), 121 | 12.3 (0.5), 121 | **<0.001** | 9.5 (0.5), 121 | 7.8 (0.4), 121 | **0.009** |
| YBOCS-SR total score | 17.4 (0.9), 112 | 16.8 (0.8), 113 | 0.563 | 12.6 (0.8), 111 | 10.9 (0.7), 113 | 0.098 |
| Exposure hierarchy completion (%) |  |  |  | 36.8 (1.9), 121 | 36.3 (1.9), 121 | 0.85 |
| Length of Stay (days) [range] |  |  |  |  |  |  |
| Inpatient |  |  |  | 33.7 (5.4), 3 (23-40) | 38.5 (18.5), 2 [20-57] | 0.775 |
| Residential |  |  |  | 63.2 (4.5), 58 [13-189] | 50.1 (2.7), 47 [14-98] | **0.022** |
| Partial hospitalization |  |  |  | 35.5 (1.5), 55 [11-70] | 31.4 (1.4), 65 [10-72] | **0.046** |
| Intensive outpatient |  |  |  | 24.4 (3.9), 5 [17-38] | 25.0 (3.4), 7 [14-42] | 0.911 |
| Readmissions (%), n) |  |  |  | 3.3, 4 | 0.0, 0 |  |

Missing values not displayed

### Clinical improvement in both groups across all measures

The anxiolytic group demonstrated a 40% reduction in ED behavior frequency and a 36% improvement in symptom severity using the EDEQ and YBC-EDS, respectively. This degree of improvement was consistent in the non-anxiolytic group, with a 46% reduction in ED behavior and a 41% improvement in symptom severity (*p* < 0.05) (**Figure 1a-b**). Both groups had similar improvement in depressive symptoms, as measured by the QIDS, with the anxiolytic group having a 36% reduction in score, and the non-anxiolytic group 37%. Co-occurring OCD symptoms, assessed with the YBOCS-SR, improved in both the anxiolytic and non-anxiolytic groups by 27% and 35%, respectively. A clinically significant change in depressive symptoms was observed for both groups, with the anxiolytic group reporting a 36% reduction in symptoms, and the non-anxiolytic group reporting a 37% reduction (*p* < 0.05) (**Figure 1c-d**). Exposure hierarchy completion was similar between the anxiolytic group (37%) and the non-anxiolytic group (36%) (**Figure 1e**). Readmission frequency did not significantly differ between treatment groups, which was 3% and 0% for the anxiolytic and the non-anxiolytic groups, respectively. Further analyses breaking down by level of care in addition to treatment status demonstrated that LOS differed at the residential and partial hospitalization levels only, with the anxiolytic patients requiring more treatment days than the non-anxiolytic patients, with no differences in LOS for inpatient and intensive outpatient levels of care (**Table 1**).

**Figure 1.**
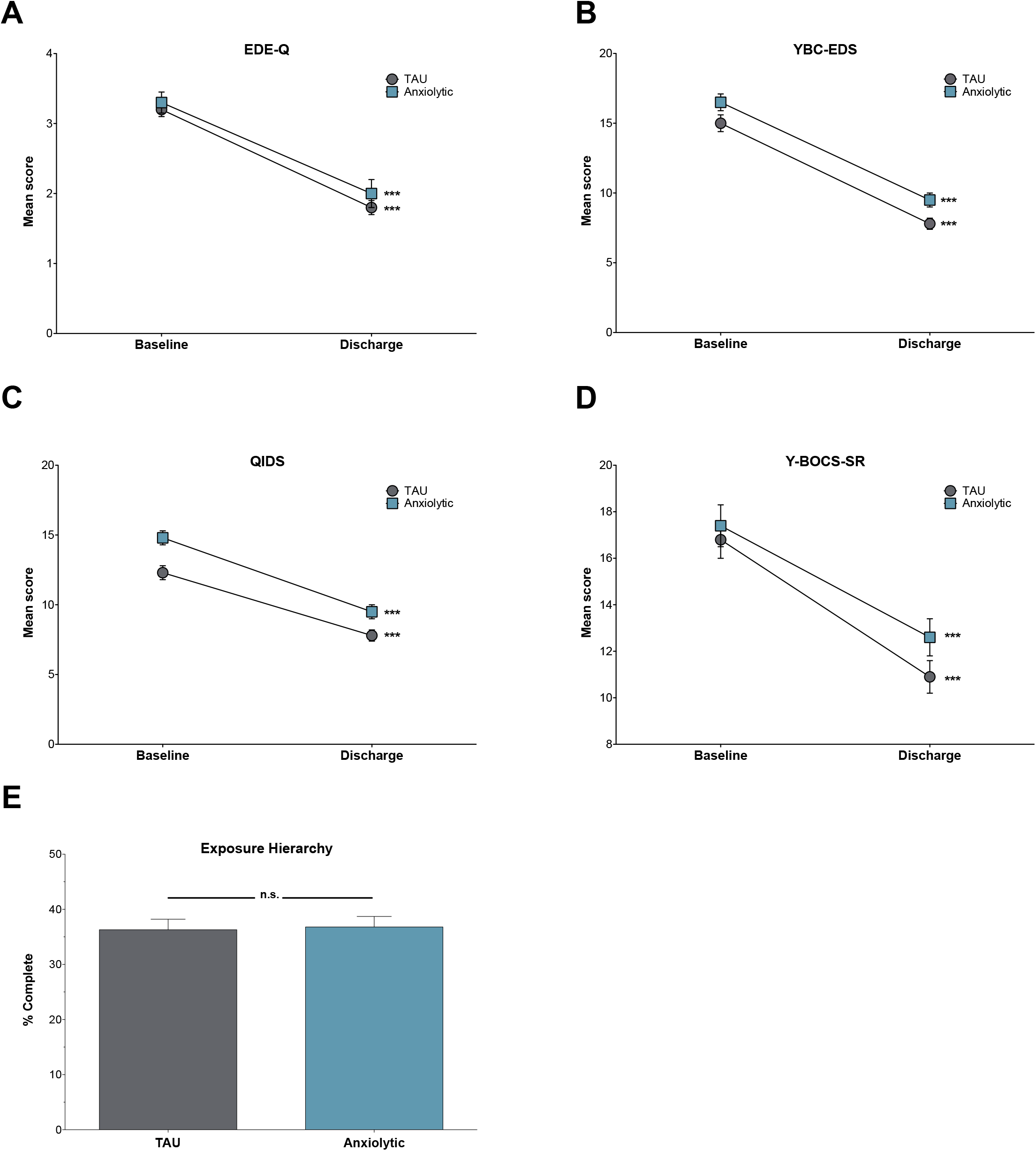
Non-anxiolytic- and anxiolytic-treated ED patients improved across all measures. (**a**) Anxiolytic-treated patients had a 40% improvement in EDE-Q score (*p* <0.001; Effect Size (ES) 0.64), and non-anxiolytic-treated patients has a 46% improvement (*p* <0.001; ES 0.72). (**b**) Anxiolytic-treated patients had a 36% improvement in YBC-EDS (*p* <0.001; ES 0.58), and non-anxiolytic-treated patients has a 41% improvement (*p* <0.001; ES 0.73). (**c**) Anxiolytic-treated patients had a 36% improvement in QIDS (*p* <0.001; ES 0.74), and non-anxiolytic-treated patients has a 37% improvement (*p* <0.001; ES 0.88). (**d**) Anxiolytic-treated patients had a 27% improvement in YBOCS (*p* <0.001; ES 0.67), and non-anxiolytic-treated patients has a 35% improvement (*p* <0.001; ES 0.90). (**e**) Non-anxiolytic-treated patients had a similar completion of their exposure hierarchy to the anxiolytic treated group (*p* <0.001).

## Discussion

This retrospective analysis of anxiolytic utilization in ED patients suggests that treatment progress with CBT was unaffected. The anxiolytic and non-anxiolytic groups were observed to be in similar clinical ranges at baseline (e.g., moderate). Importantly, both treatment groups showed marked improvement in ED symptomatology, reaching mild levels at discharge, implying that anxiolytics did not impact the rate of improvement or CBT efficacy. Furthermore, both treatment groups exhibited significant reductions in OCD and depressive symptoms from admission to discharge, indicating that patient clinical progress altogether was uninhibited by administration of anxiolytic medication (**Table 1**).

The current study, investigating a large ED patient cohort, addresses longstanding concerns of anxiolytics compromising CBT efficacy and supports that anxiolytics can be administered alongside standard therapy. The hesitancy surrounding concurrent anxiolytic use for ED patients undergoing CBT can be largely attributed to a dearth of controlled trials conducted with mixed results on this treatment combination (P. J. Hay & Claudino, 2012), particularly with high-acuity patient participants being treated in more intensive levels of care (e.g., residential, partial hospitalization). The current analysis captures an integral characteristic within this population that is not well-represented in the literature, that is, the usage of anxiolytic medication for patients with severe cognitive rigidity at baseline. This quality improvement project corroborates effectiveness of anxiolytics, particularly in early stages of treatment, when an individual may have trouble initially engaging in robust CBT due to high levels of specific and/or general anxiety.

## ETHICS DECLARATION

This Rogers Behavioral Health Institutional Review Board of Rogers Behavioral Health determined that this quality improvement project was non-human subjects research and waived ethical approval for this work

## Data availability statement

All data produced in the present study are available upon reasonable request to the authors.

