## Supplementary Table 1 for "Anxiolytic management in eating disorder patients receiving cognitive behavioral therapy: A quality improvement brief report"

| **Supplementary Table 1. Anxiolytic medications during treatment.** | |
|---|---|
| | **mean days (SEM) [range days], n** |
| Alprazolam | 36.4 (5.1) [2 – 76], 6 |
| Clonazepam | 35.2 (4.0) [1 – 112], 17 |
| Diazepam | 32.9 (8.4) [1 – 50], 2 |
| Gabapentin | 20.1 (2.0) [1 – 87], 37 |
| Hyproxyzine | 27.4 (2.2) [1 – 141], 68 |
| Lorazepam | 30.4 (3.4) [1 – 98], 21 |
